# Real-world Opioid-Sparring Effects of Infiltration between the Popliteal Artery and the Capsule of the Knee (IPACK) in Patients Undergoing Anterior Cruciate Ligament Reconstruction

**DOI:** 10.64898/2026.02.25.26346957

**Authors:** Anders Peder Højer Karlsen, Markus Harboe Olsen, Kristoffer Barfod, Troels Haxholdt Lunn, Martin Bitsch, Sebastian Wiberg, Jens Laigaard

## Abstract

**Introduction:** Patients undergoing anterior cruciate ligament (ACL) reconstruction experience substantial postoperative pain, which delays recovery and leads to both immediate and long-term opioid use. In other knee procedures, infiltration between the popliteal artery and the capsule of the posterior knee (IPACK) has demonstrated analgesic and opioid reducing effects. However, the effect in patients undergoing ACL reconstruction has not been investigated. We aimed to investigate the real-world effect of IPACK in patients undergoing ACL reconstruction on immediate postoperative opioid consumption.

**Participants:** In this single-centre difference-in-differences cohort study, all patients who underwent ACL reconstruction surgery at Bispebjerg Hospital, Denmark, from 1 February 2024 to 30 June 2025 are included. The study further includes a similar reference cohort, comprising all patients who underwent trochleaplasty, Elmslie-Trillat, or medial patellofemoral ligament reconstruction during the same period, and at the same hospital.

**Intervention:** The primary exposure is the implementation of IPACK as part of perioperative management for ACL reconstruction on 1 January 2025. The IPACK was performed under ultrasound guidance, immediately before surgery, administering 20 mL of ropivacaine 0.5% between the popliteal artery and the posterior knee capsule.

**Outcomes:** The primary outcome is the cumulative opioid consumption from surgical incision to 2 hours postoperatively. Secondary outcomes include the cumulative opioid consumption from incision to 24 hours postoperatively, the worst reported pain score at 0-24h postoperatively, occurrence of postoperative nausea or vomiting (PONV) 0-24h postoperatively, length of PACU stay, length of hospital stay, and nerve injuries. As an exploratory outcome, carbon dioxide emissions will be investigated.

**Statistical analysis:** The main analysis will be a standard two-way fixed effects DiD regression assessing the changes occurring at the time of implementation of IPACK in the ACL cohort, with adjustment for the underlying time trend. Continuous outcomes are reported as mean difference (95% confidence interval [CI]), and binary outcomes as absolute and relative risks (95% CI).

## Introduction

Patients undergoing anterior cruciate ligament (ACL) reconstruction have substantial postoperative pain, which hampers mobilisation and discharge. This delay has implications for both the patient and society, since longer hospital stays are costly. The pain is therefore often treated with fast-acting opioids, which can also hamper recovery and lead to persistent, long-term opioid use.^1^ To reduce pain and opioid consumption, multimodal analgesia is often used.^1, 2^ However, to our knowledge, the infiltration between the popliteal artery and the capsule of the posterior knee (IPACK) block has not yet been investigated in ACL reconstruction.^4 5^

The use of before-and-after study designs are susceptible to confounding by the underlying trend in the outcome over time. Interrupted time series regression can be used to evaluate the effects of an interventions by attempting to adjust for this trend,^6^ and including a reference group can further enhance the analysis, known as a difference-in-differences (DiD) analysis.^7^

Healthcare interventions should be evaluated for both patient-reported outcome measures (PROMs, e.g. pain intensity), objectively measured outcomes (e.g. opioid consumption), organisational aspects (e.g. length of stay), harms, and - where possible - economic aspects (e.g. procedural costs), and environmental outcomes (e.g. carbon emissions).^9-11^ We chose opioid consumption which is a relatively objective outcome measure, as the primary outcome, since our study is observational and non-blinded.^8^ Thus, the aim of this DiD study is to study the real-world opioid-sparring effect of IPACK in patients undergoing ACL reconstruction surgery.

## Methods

This DiD study will be reported in accordance with the Strengthening the Reporting of Observational Studies in Epidemiology (STROBE) guideline.^12^ This protocol and statistical analysis plan will be uploaded to Zenodo^13^ prior to any data inspection or analysis. We were granted ethical approval on 23 June 2025, by the Research Service Division, Capital Region of Denmark (R-25041576). The study was deemed exempt from ethical notification from the ethics committee (F-25043157; 30 June 2025). The study was approved by The Capital Region of Denmark’s Legal Department (p-2025-19423; 15 July 2025).

### Participants

We will include all patients who underwent ACL reconstruction surgery at Bispebjerg Hospital, Denmark, from 1 February 2024 to 30 June 2025, regardless of age (ACL cohort). The study further includes a similar reference cohort, comprising all patients who underwent trochleaplasty, Elmslie-Trillat, or medial patellofemoral ligament reconstruction (MPFL) during the same period, and at the same hospital (troch/Elm/MPFL cohort). All data were extracted from the electronic patient record system Sundhedsplatformen (Epic Systems Corporation, WI, USA) and cleaned using standardised protocols.^14^

### Exposure

The primary exposure is the implementation of IPACK as part of perioperative management for ACL reconstruction. The IPACK was performed under ultrasound guidance, immediately before surgery, administering 20 mL of ropivacaine 0.5% between the popliteal artery and the posterior knee capsule. The aim of IPACK is to anaesthetise the popliteal branches of the tibial, common peroneal and obturator nerves.

Patients in the ACL cohort who underwent surgery *prior* to IPACK implementation on 1 January 2025 constitute the control group, and those treated *after* serves as the intervention group. The reference cohort (troch/Elm/MPFL cohort) underwent comparable sports surgeries of the knee, performed by the same surgeons and at the same hospital, but were not subject to IPACK. The troch/Elm/MPFL cohort was included to account for unanticipated time-related changes in perioperative care.

### Anaesthesia and analgesia

Standardised perioperative care pathways were used throughout the study period in both cohorts. General anaesthesia was induced with propofol and maintained with total intravenous anesthesia using propofol and remifentanil. Patients received laryngeal masks or endotracheal intubation. Patients received pre- or intraoperative oral or intravenous paracetamol 1g, NSAIDs unless contraindicated (oral ibuprofen 400 mg or intravenous ketorolac 15-30mg), intravenous dexamethasone either 4 mg or 16 mg, and ondansetron 4 mg intravenously. Further, patients received an ultrasound guided ACB of 20 ml ropivacaine 0.5%. Postoperative analgesia consisted of opioids as needed and non-opioid adjuncts. No changes were made to the surgical, anaesthetic or analgesic standard protocol during the study period.

### Outcomes

The primary outcome is the cumulative opioid consumption from surgical incision to 2 hours postoperatively, measured in intravenous morphine equivalents (IMEs, mg).

Secondary outcomes include the cumulative opioid consumption from incision to 24 hours postoperatively or till discharged (IMEs, mg), the worst reported pain score at 0-24h postoperatively (0-10 numeric rating scale (NRS) ratings), occurrence of postoperative nausea or vomiting (PONV) within 0–24 hours, length of PACU stay (minutes), length of hospital stay (hours), and occurrence of a nerve injury that led to a hospital contact 0-90 days postoperatively.

As an exploratory outcome, we will assess the carbon emission per case only based on utensils for the IPACK procedure as cradle-to-grave CO2 equivalents. However, analysis of this outcome requires that the manufacturers of the block needle and local anaesthetic drug are willing to provide the necessary information.

## Statistical analysis

### Data handling

Opioid requirements were converted to IMEs using established conversion factors.^15^ Verbal pain ratings, although rarely used at our hospital, will be converted to 0-10 NRS values using derived values from Moore et al: No pain = 0, Mild = 1.5, Moderate = 5, Severe = 8, Unbearable pain = 10.^16^ The occurrence of PONV was treated as a binary outcome, defined as the occurrence of moderate-, severe-, or unspecified nausea, or an episode of vomiting.

### Population

The primary analysis will be the per-protocol population, i.e. including only the participants who received the intended treatment. However, we will also perform a sensitivity analysis on the intention-to-treat (ITT) population, meaning that all eligible patients are analysed regardless of actual treatment received. These analyses are included to assess the possible impact of confounding by indication bias. This bias can arise from clinicians performing IPACK in patients prone to high levels of pain and opioid use, be-fore IPACK was officially implemented. Moreover, clinicians may preclude IPACK in patients with expected low levels of pain.

### Missing data handling

We expect limited missing data. Thus, no imputation will be performed, meaning that all analyses will be conducted using complete case datasets. Missing data will be reported for each variable and outcome, and the number of observations included in each analysis will be stated explicitly.

### Main analysis

The main analysis will be a standard two-way fixed effects DiD regression, with time included as a continuous variable. This simple approach was chosen because no other changes were made to the surgical or analgesic treatment of these patients during the study period. P-values under 0.05 are considered statistically significant.

The model for the **continuous outcomes** will look like this in R:

*DiD_model <- lm(outcome ∼ surgery + surgery:after_IPACK_ + time2, data = d)*

Where **surgery** indicate the surgical cohort (ACL or troch/Elm/MPFL), **after_IPACK** is a binary indicator of IPACK implementation, and **time2** is a time variable which resets at the time of IPACK implementation, i.e. as *days since study start* in the period before implementation and *days since implementation* in the period after implementation. We chose to reset the time variable at time of implementation to reduce collinearity. The *surgery:pre_post* interaction is the DiD (average treatment effect) estimate, which will be reported with 95% confidence intervals.

For **binary outcomes**, we will report both the absolute risk difference (RD) and the relative risk (RR). The RD is estimated with a linear probability model (LPM) and heteroskedasticity-consistent and small-sample-corrected 95% confidence intervals:

*library(sandwich); library(lmtest)*

*DiD_RD <- lm(binary_outcome ∼ surgery + surgery:after_IPACK_ + time2, data = d)*

*coeftest(DiD_RD, vcov = vcovHC(DiD_RD, type = “HC1”))*

A modified poisson regression will be used to estimate the relative risk (RR):

*DiD_RR <- glm(binary_outcome ∼ surgery + surgery:after_IPACK_ + time2*,

*data = d*,

*family = poisson(link = “log”))*

*coeftest_RR <- coeftest(DiD_RR, vcov = vcovHC(DiD_RR, type = “HC1”))*

### Sensitivity analyses

#### No within-surgeon correlation assumption

Patients treated by the same surgeon may have more similar outcomes (e.g., due to surgeon technique, opioid prescribing habits, or patient selection). In the main analysis, we assume that outcomes for patients treated by the same surgeon are independent, because clustering may reduce statistical power due to the smaller effective sample size. To evaluate the possible impact of this clustering, we will conduct a sensitivity analysis with standard errors clustered at the surgeon level:

*library(sandwich); library(lmtest)*

*clustered_se <- vcovCL(DiD_model*,

*cluster = ∼surgeon_id*

*data = d)*

*coeftest(DiD_model, vcov = clustered_se)*

Where *DiD_model* is the model used in the main analysis and *surgeon_id* is a categorical (factor) variable with surgeon identifiers

#### Parallel trends between groups assumption

The main analysis assumes that, in the absence of the IPACK implementation, the outcome trends for the ACL cohort (treated) and troch/Elm/MPFL cohort (control) would have evolved in parallel over the entire study period. To evaluate for potential violations of this assumption, we will add an interaction between surgery group and the time variable (*surgery:time2*), to allow for group-specific linear trends:

*DiD_sens1 <- lm(outcome ∼ surgery + surgery:after_IPACK_ + time2 + surgery:time2, data = d)*

We will assess if the surgery:time2 interaction coefficient is statistically significant (P<0.05), suggesting a violation of the parallel trends assumption, and compare the DiD estimate and model fit to the main analysis, to assess sensitivity to violations of the parallel trends assumption.

#### No learning-effects assumption

Although we do not expect an interaction effect between time and the effect of the IPACK implementation, improved technique may have caused the intervention-effect to increase over time. To evaluate this possibility, we will estimate a model allowing the slope of the outcome over time to be different before and after IPACK implementation by adding an interaction term between the time variable and the intervention variable (*after_IPACK:time2*):

*DiD_sens2 <- lm(outcome ∼ surgery + surgery:after_IPACK_+ time2 + after_IPACK_:time2, data = d)*

Again, We will assess if the *after_IPACK:time2* interaction coefficient is statistically significant (P<0.05), suggesting a violation of the no learning-effects assumption, and compare the DiD estimate and model fit to the main analysis, to assess sensitivity to violations of the no learning-effects assumption.

#### Analysis of time trends

To explore the dynamic effects of IPACK implementation over time and further assess the parallel trends assumption, we will apply statistical process control with control charts to the ACL cohort and the troch/Elm/MPFL separately using the qicharts2 package in R.

We will apply the two ‘Anhøj rules’ and Shewhart’s 3-sigma rule to detect if a significant change occurred in each outcome during the study period, and specifically if a shift happened at the time of IPACK implementation.^17^ Calendar time will be discretized into two-months intervals relative to IPACK implementation, yielding 6 pre-implementation and 6 post-implementation periods to ensure enough time points for valid charts albeit avoiding extensive granularity. With the anhøj rules, the control charts are evaluated based on the distribution of data points in relation to the mean: unusually long runs (consecutive data points higher/lower than the mean) or unusually few crossings over the mean.

We will use ‘Xbar’ control charts for continuous outcomes and ‘P’ charts for binary outcomes. The control charts will be presented graphically (Figure 2). The statistical process control analyses are intended only to visualise the data and support the assessment of model assumptions.

## Tables

<u>Table 1:</u> The per-protocol population’s demographics, including age, sex, key comorbidities, length of surgery, type of surgery etc., and stratified by group allocation, i.e. to enable comparisons between the IPACK group and the control group (pre-IPACK group).

<u>Table S1:</u> The intention-to-treat population’s demographics, including age, sex, key comorbidities, length of surgery, type of surgery etc., and stratified by group allocation, i.e. to enable comparisons between the IPACK group and the control group (pre-IPACK group).

## Figures

<u>Figure S1</u> will be a flow diagram of the troch/Elm/MPFL cohort participants’ flow through the study, similar to Figure 1.

<u>Figure 2</u> will be control charts of the primary and secondary outcomes over time for the ACL and control cohorts.

<u>Figure 3</u> will be the distribution of worst pain scores, in the first 24 hours after ACL reconstruction, in the before IPACK-arm and the after IPACK-arm.

## Results TEMPLATE

A total of XX patients were included in the primary per-protocol analysis: XX patients in the IPACK group, XX in the control group (Figure 1), and XX in the troch/Elm/MPFL reference group (Figure S1). Baseline characteristics including age, sex, ASA classification, and number of SKS-coded procedures were XX between the IPACK group and the control group (Table 1). The median age was XX years [IQR XX – XX], and XX % were female. XX changes in any primary or secondary outcomes were detected in the reference cohort throughout the study period.

**Figure 1.**
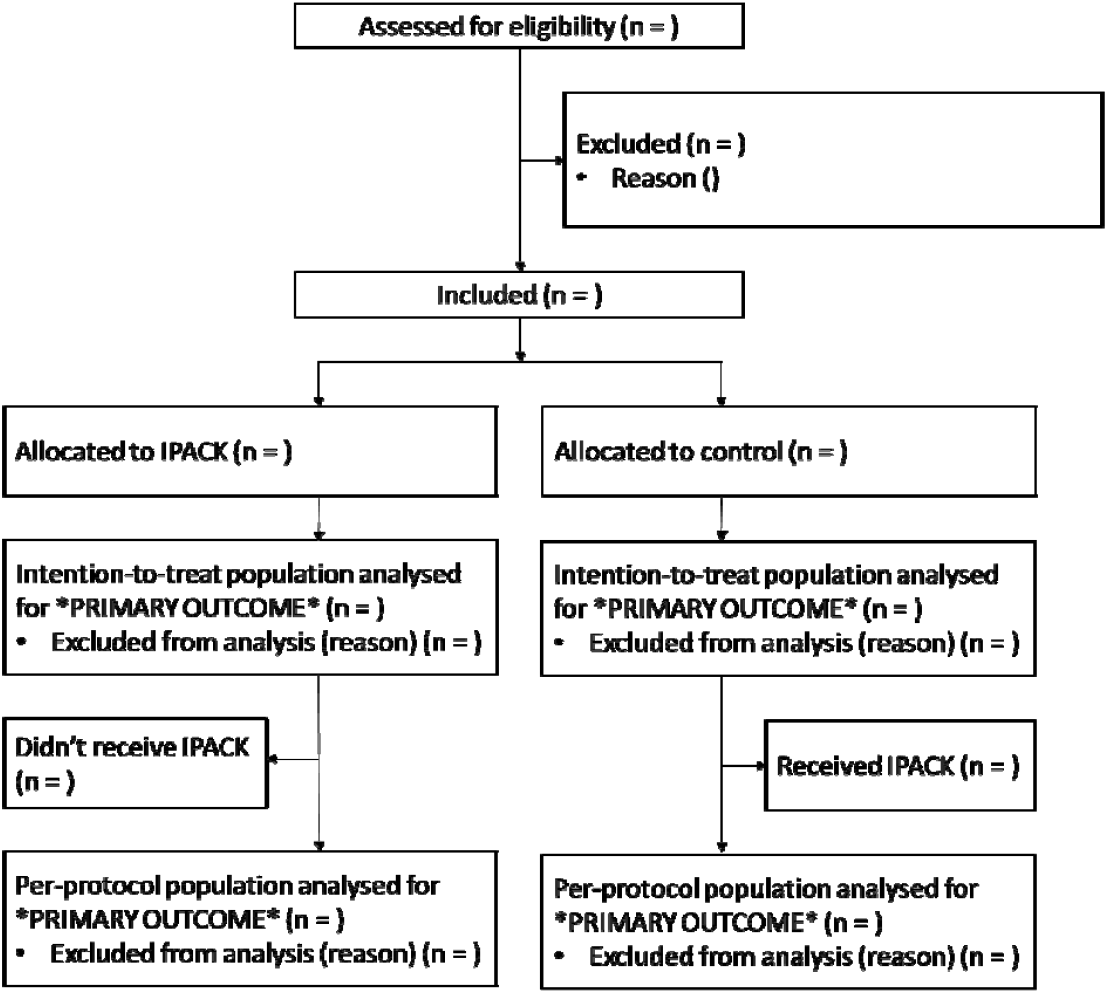
IPACK cohort participants’ flow through the study.

### Primary outcome

The ACL cohort consumed mean XX mg IMEs before implementation of IPACK (i.e. the control group), while the ACL cohort consumed mean XX mg IMEs after implementation (Figure 2). The DiD analysis suggested that implementation of IPACK led to a mean XX mg decrease/increase in cumulative opioid consumption from incision to 2 hours postoperatively (95% CI XX to XX; p = XX).

### Secondary outcomes

The DiD analyses suggested that XX OUTCOME improved/worsened by XX (95% CI XX to XX; p = XX) with implementation of IPACK [repeat for findings with p<0.05].

We found no significant effect of the implementation of IPACK on XX OUTCOME (95% CI XX to XX; p = XX) [repeat for findings with p>0.05].

## Data availability

This is a protocol paper. All data produced in the present work are contained in the manuscript.

## References

1 Fernandez CF, Angeles BS. Comparison of Analgesic Efficiency between Local Infiltration of a Long-Acting Analgesic and Regional Nerve Block among Patients Undergoing Arthroscopic Anterior Cruciate Ligament Reconstruction: Meta-Analysis of Randomized Controlled Trials. Clin Orthop Surg 2025; 17: 228–37

2 Lynch JR, Okoroha KR, Lizzio V, Yu CC, Jildeh TR, Moutzouros V. Adductor Canal Block Versus Femoral Nerve Block for Pain Control After Anterior Cruciate Ligament Reconstruction: A Prospective Randomized Trial. Am J Sports Med 2019; 47: 355–63

3 Hussain N, Brull R, Sheehy B, Dasu M, Weaver T, Abdallah FW. Does the addition of iPACK to adductor canal block in the presence or absence of periarticular local anesthetic infiltration improve analgesic and functional outcomes following total knee arthroplasty? A systematic review and meta-analysis. Reg Anesth Pain Med 2021; 46: 713–21

4 Hussain N, Brull R, Vannabouathong C, et al. Network meta-analysis of the analgesic effectiveness of regional anaesthesia techniques for anterior cruciate ligament reconstruction. Anaesthesia 2023; 78: 207–24

5 Rao N, Triana J, Avila A, et al. Postoperative Pain and Opioid Usage With Combined Adductor Canal and IPACK Block Versus Isolated Adductor Canal Block After Anterior Cruciate Ligament Reconstruction With a Bone-Patellar Tendon-Bone Autograft: A Single-Center Randomized Controlled Trial. Am J Sports Med 2025; 53: 1359–67

6 Bernal JL, Cummins S, Gasparrini A. Interrupted time series regression for the evaluation of public health interventions: a tutorial. Int J Epidemiol 2017; 46: 348–55

7 Dimick JB, Ryan AM. Methods for evaluating changes in health care policy: the difference-in-differences approach. JAMA 2014; 312: 2401–2

8 Karlsen APH, Pedersen C, Laigaard J, Mathiesen O. Opioid consumption as a core outcome domain in postoperative pain management. Pain 2022; 163: e786–e7

9 Petersen JJ, Hemberg L, Thabane L, et al. Integrating environmental outcomes in randomised clinical trials: a call to action. Lancet 2025; 405: 446–8

10 Pogatzki-Zahn EM, De Lucia S, Weinmann C, et al. A core outcome set of measurement instruments for assessing effectiveness and efficacy of perioperative pain management: results of the international IMI-PainCare PROMPT Delphi consensus process. Br J Anaesth 2025; 134: 1460–73

11 Bojic S, Ladjevic N, Palibrk I, et al. Cost-effectiveness of the Perioperative Pain Management Bundle a registry-based study. Front Public Health 2023; 11: 1157484

12 von Elm E, Altman DG, Egger M, et al. Strengthening the Reporting of Observational Studies in Epidemiology (STROBE) statement: guidelines for reporting observational studies. BMJ 2007; 335: 806–8

13 Zenodo. European Organization For Nuclear Research AND OpenAIRE 2013. Available from https://zenodo.org/

14 Karlsen APH, Moller MO, Pedersen NK, et al. The AI and Automation in Anaesthesia (TRIPLE-A) Perioperative Database in Eastern Denmark 2017-: A Platform for Epidemiology, AI-Driven Prediction, Quality Control and Automated Data Retrieval. Acta Anaesthesiol Scand 2025; 69: e70116

15 Tran TXM, Wetterslev M, Norskov AK, et al. Intraoperative opioid administrations, rescue doses in the post-anaesthesia care unit and clinician-perceived factors for dose adjustments in adults: A Danish nationwide survey. Acta Anaesthesiol Scand 2025; 69: e70000

16 Moore RA, Clephas PRD, Straube S, Wertli MM, Ireson-Paige J, Heesen M. Comparing pain intensity rating scales in acute postoperative pain: boundary values and category disagreements. Anaesthesia 2024; 79: 139–46

17 Anhoj J, Olesen AV. Run charts revisited: a simulation study of run chart rules for detection of non-random variation in health care processes. PLoS One 2014; 9: e113825

