## Supplementary material for "Real-world Opioid-Sparring Effects of Infiltration between the Popliteal Artery and the Capsule of the Knee (IPACK) in Patients Undergoing Anterior Cruciate Ligament Reconstruction": R code

### Sham data analysis - IPACK Difference-in-differences study

Jens Laigaard

2025-10-10

This documents describes the analysis for the IPACK difference-in-differences study. Note that the data and results are completely arbitrary (and does not reflect the authors' beliefs or hypotheses).

#### Load packages

```
library(tidyverse)
```

### Create sham data

```
set.seed(1234567)

n <- 400  # Number of observations

# Base variables
d <- data.frame(
  time = sample(1:730, n, replace = TRUE),  # Days since study start
  surgery = sample(c("troch/Elm/MPFL", "ACL"), n, replace = TRUE, prob = c(0.6, 0.4)),
  surgeon_id = sample(paste0("S", 1:20), n, replace = TRUE),  # Categorical surgeon ID
  outcome <- rnorm(n, mean = 10, sd = 2) # continuous outcome
)

# Create before/after variable
d <- d %>%
  mutate(after_IPACK_ = if_else(time < 365, 0, 1)) %>%
  mutate(time2 = if_else(time >= 365, time-365, time))


# Change troch/Elm/MPFL outcomes for all time points (baseline difference)
d <- d %>%
  mutate(outcome = ifelse(surgery == "troch/Elm/MPFL", outcome * 1.5, outcome))


# Reduce ACL outcomes by 4 mg only in the post-IPACK-implementation period (intervention effe
ct)
d <- d %>%
  mutate(outcome = ifelse(surgery == "ACL" & after_IPACK_ == 1, outcome-4, outcome))

# Binary outcome
d$prob <- 0.4  # Default probability
d$prob[d$surgery == "ACL" & d$after_IPACK_ == 1] <- 0.2  # Override for ACL after IPACK imple
mentation
d$binary_outcome <- rbinom(nrow(d), 1, d$prob) # Make binary


# Basic scatterchart
ggplot(d, aes(x = time, y = outcome, color = surgery)) +
  geom_point(size = 3, alpha = 0.6) +
  geom_vline(xintercept = 365, linetype = "dashed", color = "black", alpha = 0.5) +
  theme_minimal()
```

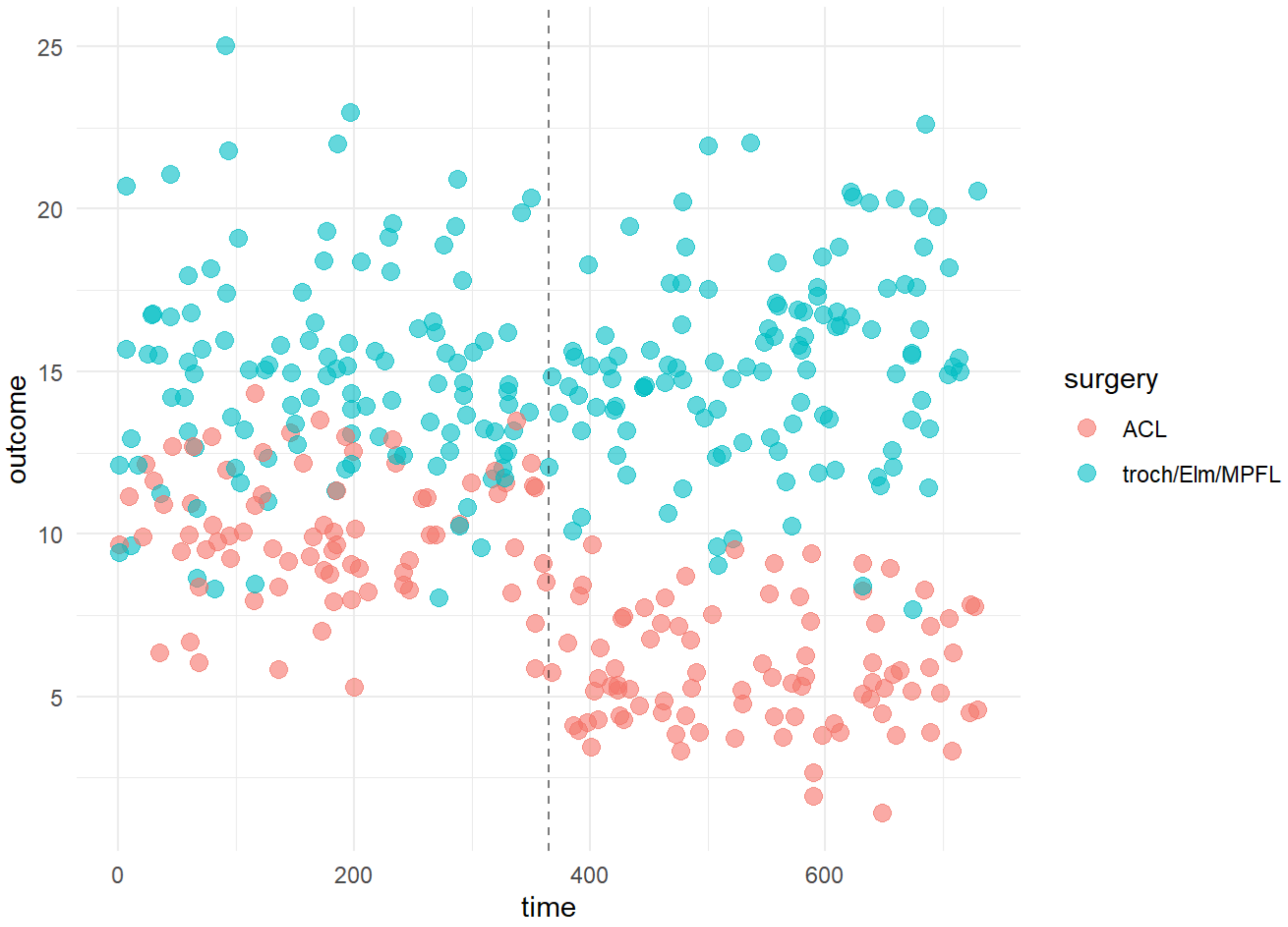


```
# View binary outcome data
table(d$binary_outcome, d$surgery, d$after_IPACK_)
```

```
## , ,  = 0
##
##
##      ACL troch/Elm/MPFL
##   0  44             68
##   1  35             47
##
## , ,  = 1
##
##
##      ACL troch/Elm/MPFL
##   0  72             72
##   1  17             45
```

---

### CONTINUOUS OUTCOMES: Main analysis

---

```
# step by step modelling:

# t test
ttest_model <- lm(outcome ~ after_IPACK_,
                  data = d[d$surgery=="ACL",])
summary(ttest_model) # Simple t-test shows 4 mg higher use i ACL without IPACK (pre) vs. ACL+
IPACK (post)
```

```
##
## Call:
## lm(formula = outcome ~ after_IPACK_, data = d[d$surgery == "ACL",
##     ])
##
## Residuals:
##     Min      1Q  Median      3Q     Max
## -4.7662 -1.3163 -0.1764  1.4966  4.2755
##
## Coefficients:
##              Estimate Std. Error t value Pr(>|t|)
## (Intercept)   10.0627     0.2143   46.96   <2e-16 ***
## after_IPACK_  -4.2827     0.2944  -14.55   <2e-16 ***
## ---
## Signif. codes:  0 '***' 0.001 '**' 0.01 '*' 0.05 '.' 0.1 ' ' 1
##
## Residual standard error: 1.904 on 166 degrees of freedom
## Multiple R-squared:  0.5604, Adjusted R-squared:  0.5578
## F-statistic: 211.6 on 1 and 166 DF,  p-value: < 2.2e-16
```

```
# Adjusted for time (underlying trend = interrupted time series analysis)
ITS_model <- lm(outcome ~ after_IPACK_ + time2,
                data = d[d$surgery=="ACL",])
summary(ITS_model) # NB in this sham data, there is no time effect - thus adding 'time' only
removes power due to co-linearity
```

```
##
## Call:
## lm(formula = outcome ~ after_IPACK_ + time2, data = d[d$surgery ==
##     "ACL", ])
##
## Residuals:
##     Min      1Q  Median      3Q     Max
## -4.7600 -1.3158 -0.1913  1.5243  4.2582
##
## Coefficients:
##               Estimate Std. Error t value Pr(>|t|)
## (Intercept)  10.1122311  0.3305547  30.592   <2e-16 ***
## after_IPACK_ -4.2829358  0.2952484 -14.506   <2e-16 ***
## time2        -0.0002786  0.0014115  -0.197    0.844
## ---
## Signif. codes:  0 '***' 0.001 '**' 0.01 '*' 0.05 '.' 0.1 ' ' 1
##
## Residual standard error: 1.91 on 165 degrees of freedom
## Multiple R-squared:  0.5605, Adjusted R-squared:  0.5552
## F-statistic: 105.2 on 2 and 165 DF,  p-value: < 2.2e-16
```

```
# Adding type of surgery and interaction between intervention and surgery. There should be no effect in the other surgical group
DiD_model <- lm(outcome ~ surgery + surgery:after_IPACK_ + time2,
                data = d)
summary(DiD_model) # See the surgeryACL:after_IPACK_ term.
```

```
##
## Call:
## lm(formula = outcome ~ surgery + surgery:after_IPACK_ + time2,
##     data = d)
##
## Residuals:
##     Min      1Q  Median      3Q     Max
## -7.6973 -1.6402 -0.1914  1.5009 10.3295
##
## Coefficients:
##                                   Estimate Std. Error t value Pr(>|t|)
## (Intercept)                       9.791233   0.377686  25.924   <2e-16 ***
## surgerytroch/Elm/MPFL             4.764171   0.389399  12.235   <2e-16 ***
## time2                             0.001525   0.001291   1.181    0.238
## surgeryACL:after_IPACK_          -4.281446   0.411877 -10.395   <2e-16 ***
## surgerytroch/Elm/MPFL:after_IPACK_  0.355033   0.349883   1.015    0.311
## ---
## Signif. codes:  0 '***' 0.001 '**' 0.01 '*' 0.05 '.' 0.1 ' ' 1
##
## Residual standard error: 2.665 on 395 degrees of freedom
## Multiple R-squared:  0.6764, Adjusted R-squared:  0.6731
## F-statistic: 206.4 on 4 and 395 DF,  p-value: < 2.2e-16
```

```
# IPACK improved (reduced) the outcome more in the ACL arm than in the troch/Elm/MPFL arm (be
cause IPACK was not introduced in the troch/Elm/MPFL arm)
```

### CONTINUOUS OUTCOMES: Sensititivy analyses

```
# --------------------------------------------
# --- Within-surgeon Correlation Assumption ---
# --------------------------------------------
library(sandwich); library(lmtest)
```

```
## Warning: pakke 'lmtest' blev bygget under R version 4.2.3
```

```
## Indlæser krævet pakke: zoo
```

```
## Warning: pakke 'zoo' blev bygget under R version 4.2.3
```

```
##
## Vedhæfter pakke: 'zoo'
```

```
## De følgende objekter er maskerede fra 'package:base':
##
##     as.Date, as.Date.numeric
```

```
# Recalculate CI from the main analysis based on standard errors clustered at the surgeon lev
el:
clustered_se <- vcovCL(DiD_model,
                       cluster = ~surgeon_id,
                       data = d)
coeftest(DiD_model, vcov = clustered_se)  # Coefficients with clustered SEs
```

```
##
## t test of coefficients:
##
##                                       Estimate Std. Error  t value Pr(>|t|)
## (Intercept)                          9.7912325  0.2977494  32.8841   <2e-16 ***
## surgerytroch/Elm/MPFL               4.7641712  0.3888708  12.2513   <2e-16 ***
## time2                                0.0015253  0.0012774   1.1940   0.2332
## surgeryACL:after_IPACK_            -4.2814463  0.3167147 -13.5183   <2e-16 ***
## surgerytroch/Elm/MPFL:after_IPACK_  0.3550334  0.4067557   0.8728   0.3833
## ---
## Signif. codes:  0 '***' 0.001 '**' 0.01 '*' 0.05 '.' 0.1 ' ' 1
```

```
# ------------------------------------------------
# --- Parallel trends between groups assumption ---
# ------------------------------------------------
# Add a surgery:time interaction, allowing for different underlying time trends between surgi
cal cohorts
DiD_model_sens1 <- lm(outcome ~ surgery + surgery:after_IPACK_ + time2 + surgery:time2,
                data = d)
summary(DiD_model_sens1)
```

```
##
## Call:
## lm(formula = outcome ~ surgery + surgery:after_IPACK_ + time2 +
##     surgery:time2, data = d)
##
## Residuals:
##    Min     1Q Median     3Q    Max
## -7.869 -1.666 -0.187  1.544 10.453
##
## Coefficients:
##                                    Estimate Std. Error t value Pr(>|t|)
## (Intercept)                      10.1122311  0.4608556  21.942  < 2e-16 ***
## surgerytroch/Elm/MPFL             4.1958520  0.6087637   6.892 2.19e-11 ***
## time2                            -0.0002786  0.0019679  -0.142    0.887
## surgeryACL:after_IPACK_          -4.2829358  0.4116321 -10.405  < 2e-16 ***
## surgerytroch/Elm/MPFL:after_IPACK_  0.3541809  0.3496744   1.013    0.312
## surgerytroch/Elm/MPFL:time2       0.0031640  0.0026062   1.214    0.225
## ---
## Signif. codes:  0 '***' 0.001 '**' 0.01 '*' 0.05 '.' 0.1 ' ' 1
##
## Residual standard error: 2.663 on 394 degrees of freedom
## Multiple R-squared:  0.6776, Adjusted R-squared:  0.6735
## F-statistic: 165.6 on 5 and 394 DF,  p-value: < 2.2e-16
```

```
# --------------------------------------
# --- No learning effects assumption ---
# --------------------------------------
# Add a intervention:time interaction, allowing the slope of the outcome over time to be diff
erent before and after IPACK implementation
DiD_model_sens2 <- lm(outcome ~ surgery + surgery:after_IPACK_ + time2 + after_IPACK_:time2,
                data = d)
summary(DiD_model_sens2)
```

```
##
## Call:
## lm(formula = outcome ~ surgery + surgery:after_IPACK_ + time2 +
##     after_IPACK_:time2, data = d)
##
## Residuals:
##     Min      1Q  Median      3Q     Max
## -7.9152 -1.6567 -0.1557  1.5081 10.1621
##
## Coefficients:
##                                      Estimate Std. Error t value Pr(>|t|)
## (Intercept)                        10.1191930  0.4457425  22.702  < 2e-16 ***
## surgerytroch/Elm/MPFL               4.7713321  0.3889863  12.266  < 2e-16 ***
## time2                              -0.0003177  0.0018555  -0.171    0.864
## surgeryACL:after_IPACK_            -4.9143577  0.6157750  -7.981  1.6e-14 ***
## surgerytroch/Elm/MPFL:after_IPACK_ -0.2942291  0.5857044  -0.502    0.616
## after_IPACK_:time2                  0.0035647  0.0025805   1.381    0.168
## ---
## Signif. codes:  0 '***' 0.001 '**' 0.01 '*' 0.05 '.' 0.1 ' ' 1
##
## Residual standard error: 2.661 on 394 degrees of freedom
## Multiple R-squared:  0.678,  Adjusted R-squared:  0.6739
## F-statistic: 165.9 on 5 and 394 DF,  p-value: < 2.2e-16
```

---

### BINARY OUTCOMES - Main analysis

---

```
library(sandwich); library(lmtest)



# -------------------------------------
# --- Absolute Risk Difference (RD) ---
# -------------------------------------
DiD_RD <- lm(binary_outcome ~ surgery + surgery:after_IPACK_ + time2, # Linear Probability Model (LPM)
             data = d)
coeftest_RD <- coeftest(DiD_RD, vcov = vcovHC(DiD_RD, type = "HC1")) # Heteroskedasticity-consistent standard errors with small-sample correction)

# Extract the DiD estimate and SE, and calculate 95% CI
RD_estimate <- coef(DiD_RD)["surgeryACL:after_IPACK_"]
RD_se <- coeftest_RD[rownames(coeftest_RD) == "surgeryACL:after_IPACK_", "Std. Error"]

# Calculate RD 95% CI
RD_CI <- RD_estimate + c(-1.96, 1.96) * RD_se

#Print
cat("Absolute Risk Difference (RD):", round(RD_estimate, 3), "\n"); cat("95% CI:", round(RD_CI, 3), "\n")
```

```
## Absolute Risk Difference (RD): -0.252
```

```
## 95% CI: -0.389 -0.114
```

```
# ---------------------------------------
# --- Relative Risk / Risk Ratio (RR) ---
# ---------------------------------------
DiD_RR <- glm(binary_outcome ~ surgery + surgery:after_IPACK_ + time2,, # Modified Poisson regression
              data = d,
              family = poisson(link = "log"))
coeftest_RR <- coeftest(DiD_RR, vcov = vcovHC(DiD_RR, type = "HC1")) # Heteroskedasticity-consistent standard errors with small-sample correction

# Extract log(RR) estimate and SE
logRR_estimate <- coef(DiD_RR)["surgeryACL:after_IPACK_"]
logRR_se <- coeftest_RR[rownames(coeftest_RR) == "surgeryACL:after_IPACK_", "Std. Error"]

# Calculate RR and 95% CI
RR_estimate <- exp(logRR_estimate)
RR_CI <- exp(logRR_estimate + c(-1.96, 1.96) * logRR_se)

#Print
cat("Relative Risk (RR):", round(RR_estimate, 3), "\n"); cat("95% CI:", round(RR_CI, 3), "\n")
```

```
## Relative Risk (RR): 0.431
```

```
## 95% CI: 0.262 0.709
```

---

### Statistical Process Control —

---

```
library(qicharts2)

d_qi <- d %>%
  mutate(bin_2month = as.numeric(
    cut(time,
        breaks =  seq(0, 791, by = 61),
        labels = c("-12 to -10 months", "-10 to -8 months", "-8 to -6 months", "-6 to -4 mont
hs", "-4 to -2 months", "-2 to 0 months", "0 to 2 months", "2 to 4 months", "4 to 86 months",
"6 to 8 months", "8 to 10 months", "10 to 12 months"),
        include.lowest = TRUE)) - 6) # NB numeric is needed to apply

# ----------------------------------
# Continuous outcomes: Xbar Charts
# ----------------------------------
qic(bin_2month, outcome,
    data  = d_qi[d$surgery=="ACL",],
    chart = 'xbar',
    title = 'Average continuous outcome (Xbar chart) in the ACL cohort',
    ylab  = 'Outcome',
    xlab  = 'Time (2 month bins)')

qic(bin_2month, outcome,
    data  = d_qi[d$surgery=="troch/Elm/MPFL",],
    chart = 'xbar',
    title = 'Average continuous outcome (Xbar chart) in the troch/Elm/MPFL cohort',
    ylab  = 'Outcome',
    xlab  = 'Time (2 month bins)')



# ------------------
# Binary outcomes: p charts
# --- ACL cohort ---
# ------------------

d_qi_ACL <- d_qi[d_qi$surgery=="ACL",] %>% # Calculate 2-month bin counts in the ACL group
  group_by(bin_2month) %>%
  summarise(binary_outcome_count = sum(binary_outcome),
          n = n())

qic(bin_2month, binary_outcome_count, n,
    data     = d_qi_ACL,
    chart    = 'p',
    title    = 'Binary outcome (p chart) in the ACL cohort',
    ylab     = 's',
    xlab     = 'Time (2 month bins)')

# ----------------------------
# --- troch/Elm/MPFL cohort ---
# ----------------------------

d_qi_trochElmMPFL <- d_qi[d_qi$surgery=="troch/Elm/MPFL",] %>% # Calculate 2-month bin counts
in the troch/Elm/MPFL group
  group_by(bin_2month) %>%
  summarise(binary_outcome_count = sum(binary_outcome),
          n = n())
```

```
qic(bin_2month, binary_outcome_count, n,
    data     = d_qi_trochElmMPFL,
    chart    = 'p',
    title    = 'Binary outcome (p chart) in the troch/Elm/MPFL cohort',
    ylab     = 's',
    xlab     = 'Time (2 month bins)')
```
